# *OpenCodeCounts*: An open-access, interactive online tool and R package for analysing clinical code usage in England

**DOI:** 10.1101/2025.10.14.25338005

**Authors:** Arina A Tamborska, Rose Higgins, Yamina Boukari, Viveck Kingsley, Lola Ojedele, Kunle Oreagba, Jon Massey, Andrea Schaffer, Amelia Green, William Hulme, Brian MacKenna, Helen J Curtis, Louis Fisher, Milan Wiedemann

## Abstract

Clinical codes are unique identifiers used in electronic health records to document specific information, such as diagnoses, procedures or medications. Because of their structured and systematic nature, they are often used for research, audit and service evaluation. Knowing how frequently certain codes are recorded can be invaluable in planning such work. For example, not all events are recorded with equal frequency, so knowing the usage of specific codes helps determine research feasibility.

In England, data on the frequency of clinical code recording is openly available for three classification systems used in primary and secondary care: SNOMED CT, ICD-10 and OPCS-4. However, these valuable datasets, showing how frequently individual clinical code were used, are difficult to access and analyse in their current format, hindering their application in research and service evaluation.

We developed *OpenCodeCounts*, an interactive online tool with an accompanying R package that help users explore and analyse England’s primary and secondary care clinical coding data. Both are compatible with *OpenCodelists*.*org*: a publicly available and free-to-use platform for codelist development and sharing. This article describes the underlying datasets, the development of the *opencodecounts* R package and the interactive app, and showcases their applications for electronic health records research.

**Plain English Summary:** Electronic health records (EHR) are a summary of patients’ medical files. They are a useful resource for researchers who want to study health and healthcare use. EHR often use unique codes to record diagnoses, treatments, procedures and other clinical information. These codes help organise and standardise healthcare data. Researchers working with EHR need to know how often different codes are used to plan studies and build accurate lists of codes (called codelists) for their research. In England, data on how often these medical codes are used is publicly available as large spreadsheets, which makes it time-consuming to access and analyse.

To make this easier, we created an interactive online tool that allows researchers to explore and visualise this information easily, as well as an R package - a collection of tools written in the R programming language. The package, called *opencodecounts*, allows researchers to access and study this information. We made these tools compatible with a publicly available website for codelists preparation and sharing, called *OpenCodelists*.*org*. In this article, we describe the underlying data, explain how the tools were developed, and demonstrate how they can be used to support research using EHR.

## Introduction

In healthcare settings, interactions between patients and providers are recorded to create electronic health records (EHR), which are digital files outlining one’s medical history. They contain both free text and structured information, often recorded in the form of [clinical codes](https://digital.nhs.uk/developer/guides-and-documentation/building-healthcare-software/clinical-coding-classifications-and-terminology). Clinical codes are unique numeric or alphanumeric identifiers grouped under hierarchies of different classification systems. They allow for standardised recording of diagnoses, procedures or medications, as well as other clinical, demographic and administrative data. The information captured by clinical codes is frequently used for healthcare services billing and administration, and in direct patient care, where it provides a summary of an individual’s medical history. Clinical codes recorded across multiple patients also form a rich resource for health research, audit and service evaluation.

Research using EHR data typically uses structured information captured by clinical codes to identify the population of interest and other relevant variables, such as patient characteristics, exposures or clinical outcomes. A list of clinical codes identifying a specific variable is known as a codelist (and can be also known as a reference set or a phenotype). A carefully designed codelist, capturing accurately the desired variables, ensures study validity (Williams et al. 2017). Codelist development involves a search for inclusion terms via [code browsers](https://termbrowser.nhs.uk/), utilisation of [hierarchies](https://icd.who.int/browse10/2019/en) and [codelist builders](https://www.opencodelists.org/), application of exclusion terms and clinical reviews (Matthewman et al. 2024). OpenCodelists.org is a platform for developing and sharing codelists for EHR research. It provides an interface to build codelists based on term searches and contains a large collection of codelists capturing a wide range of potential variables. However, it does not provide feedback on how frequently the selected codes are used.

Knowing the frequency and accuracy of clinical coding can help both in codelist development and more widely in the EHR study design. First, it aids assessment of study feasibility, since events which are poorly recorded in EHR may not be suitable for EHR research. Understanding whether clinical events are accurately captured in the EHR is difficult, but can be examined by checking that code usage is broadly aligned with expectations based on other data sources or clinical expertise. In this way, one can also assess whether an existing codelist comprehensively captures the clinical activity and decide on the inclusion of any codes considered “borderline”. Finally, the national code usage data allows to detect changes in the recording trends, for example, due to code retirement or financial coding incentives (Beaney et al. 2023), which may impact the interpretation of the findings. It also serves as an independent benchmark for estimates derived from other EHR data sources.

### Overview of clinical code usage data in England

Whilst individual patient records can only be accessed via trusted research environments (Edwards et al. 2023), aggregate data on cumulative code usage is valuable for the above purposes. In England, NHS makes annual summaries of clinical coding activity in primary care (general practice) and secondary care (with the most complete data available for inpatient admissions) openly available. These summaries cover annual usage of SNOMED Clinical Terms ([SNOMED CT](https://www.england.nhs.uk/digitaltechnology/digital-primary-care/snomed-ct/)) in primary care; and International Statistical Classification of Diseases and Related Health Problems, 10th Revision (ICD-10) and OPCS Classification of Interventions and Procedures, version 4 OPCS-4) in secondary care. For each of the three classification systems, the summary provides annual usage for each code used at least once each year.

#### SNOMED CT use in general practice

[SNOMED CT](https://www.snomed.org/what-is-snomed-ct) is the world’s most comprehensive clinical terminology system assigning specific codes to clinical findings, observations, anatomical terms, disease causes, products and procedures. It contains [357,000 globally common codes](https://digital.nhs.uk/developer/guides-and-documentation/building-healthcare-software/clinical-coding-classifications-and-terminology#snomed-ct), with additional codes used in the UK for local-context screening procedures and products. Each SNOMED CT code has a description, including a semantic tag shown in parentheses, that identifies the code’s high-level category, such as disorder, finding, procedure, or observable entity, for example [Blood pressure recorded by patient at home](https://termbrowser.nhs.uk/?perspective=full&conceptId1=413153004&edition=uk-edition&release=v20250924&server=https%3A%2F%2Ftermbrowser.nhs.uk/sct-browser-api/snomed&langRefset=999001261000000100,999000691000001104) (procedure). In the UK, SNOMED CT has been used by healthcare providers and administrators in general practice (GP) since 2019, when the terminology replaced Read Codes Clinical Terms. In most cases, previously captured data [was translated](https://digital.nhs.uk/developer/guides-and-documentation/building-healthcare-software/clinical-coding-classifications-and-terminology#read-codes-retired) to the equivalent SNOMED CT codes.

The annual code usage is published every October by NHS England, covering the preceding August to July. The number of individuals included [has been increasing](https://files.digital.nhs.uk/D8/F9D898/SNOMED_code_usage_metadata.xlsx), reaching over 62 million patient records across 6,600 providers in 2023/24 (compared with almost 56 million in 2018/19). The annual usage count reflects how many times each listed SNOMED code was added to a GP patient record in England in a given year. The counts are rounded to the nearest 10, while counts between 1 and 4 are withheld. The codes with no usage are excluded.

#### ICD-10 and OPCS-4 use in hospital admitted patient care

The ICD-10 and OPCS-4 classification systems are used for coding diagnoses and procedures in secondary care in acute hospitals in England. ICD-10 is a global system, containing 18,000 diagnoses coded as four or five-character alphanumeric codes. OPCS-4, consisting of 11,500 four-letter alphanumeric codes, was developed by NHS Digital for recording procedures. Both are hierarchical systems, grouped in chapters corresponding to clinical areas.

ICD-10 and OPCS-4 coding determine financial reimbursement for admitted patient care in all NHS-commissioned acute hospitals in England. This results in [systematic and near-complete coding](https://digital.nhs.uk/data-and-information/data-tools-and-services/data-services/hospital-episode-statistics/the-processing-cycle-and-hes-data-quality). The coding data submitted by the hospitals is aggregated and published openly by NHS England as the Admitted Patient Care Activity of the Hospital Episode Statistics (HES-APC). It is released in annual intervals, covering April to March, in line with the NHS financial year.

Activity in HES-APC is captured in episodes of care under one consultant, known as Finished Consultant Episodes (FCEs). Each FCE can be assigned up to 20 clinical diagnoses (ICD-10 codes) and up to 24 procedures (OPCS-4 codes), with only one diagnosis and only one procedure being recorded as “primary” (Herbert et al. 2017). The usage count for ICD-10 codes equals the number of FCEs with a specific ICD-10 code recorded in any primary or secondary position. The usage count for OPCS-4 codes equals the total number of procedures recorded in primary or secondary positions across all FCEs. This means that each specific diagnosis is counted just once within an episode of care, while a procedure can be carried out (and counted) multiple times within a single episode. In other words, the count of procedures can be greater than the count of all episodes where that procedure is recorded. Importantly, until 2012/13, only ICD-10 codes used in the primary diagnosis position [were made publicly available](https://files.digital.nhs.uk/publicationimport/pub12xxx/pub12566/hosp-epis-stat-pub-review-2013-14.pdf). Also that year, a methodological change (counting each ICD-10 code only once per episode) was introduced, meaning that the preceeding counts, even if made available, would not have been directly comparable. The usage count in the HES-APC is not rounded, and the codes with no usage are excluded. Small usage counts for sensitive codes (such as non-spontaneous abortions) are suppressed. The counts are stratified by sex and age group.

### Why is this tool needed?

The national aggregate code usage statistics provide invaluable insights into coding trends, but cannot be easily used to support codelist development and other research applications. They are split across multiple datasets and published in different files with varying URL formats and different structures and formatting. To generate annual trends, the data has to be fetched, processed, systematised and aggregated. This is not a trivial task (Bacon & Goldacre 2020) and has not been achieved comprehensively. Previous studies using code usage data conducted time-limited and disease-specific analyses of coding patterns in publicly available data, often using one coding system (Zghebi et al. 2022). A centralised, well-structured, routinely updated, and easily accessible resource for all code usage datasets can significantly enhance codelist development, facilitate coding frequency analysis, and support various EHR research applications.

We therefore introduce *OpenCodeCounts*, which includes an interactive online tool and an accompanying R package, designed to facilitate the exploration and analysis of clinical code usage data in England. The online tool allows users to visualise and explore coding trends interactively, without requiring any programming experience, while the R package enables detailed and reproducible analyses. Together, these resources address the limitations of existing data by (1) providing harmonised access to all available years in a unified structure and (2) facilitating seamless integration with OpenCodelists.org, a publicly available platform with almost 2000 codelists, to streamline codelist development and other research applications.

## Methods

### Implementation

#### Data preparation

We obtained publicly available clinical code usage data from NHS England starting from 2011. The SNOMED CT datasets were available in text files, which we processed to ensure a standardised structure and correct format of the SNOMED codes. Some code descriptions in years 2018-21 contained [encoding issues](https://github.com/bennettoxford/opencodecounts/issues/39), which we corrected during preprocessing. We added the exact start (1st August) and end (31st July) dates and the appropriate year. Where code usage was suppressed due to small counts (1 - 4), we imputed a value of 5. All other SNOMED counts are rounded to the nearest 10 in the raw data, but imputing 10 would artificially inflate the cumulative usage for rarely used codes over a long period.

The ICD-10 and OPCS-4 datasets were available in Microsoft Excel format. We selected the usage counts for the four-character ICD-10 codes and the OPCS-4 codes recorded in any primary or secondary position. We added 1st April and 31st March as start and end dates, and the appropriate year. To account for the reporting changes described above, we used data from 2012 onwards. We standardised the codes to remove inconsistencies, non-alphanumeric characters and language encoding errors similar to those described above (which affected ICD-10 year 2022). In the source ICD-10 and OPCS-4 datasets, the small counts are suppressed only for codes considered sensitive, such as [non-spontaneous abortions](https://www.gov.uk/government/statistics/abortion-statistics-for-england-and-wales-2022/guide-to-abortion-statistics-england-and-wales-2022). For consistency across the datasets, we imputed a value of 5. To minimise package size, we did not incorporate breakdowns by age and gender. While these variables are informative, they increase the dataset size, making the package less efficient for general use. In the current version of the package, the columns available for each dataset are presented in Table 1.

**Table 1.**
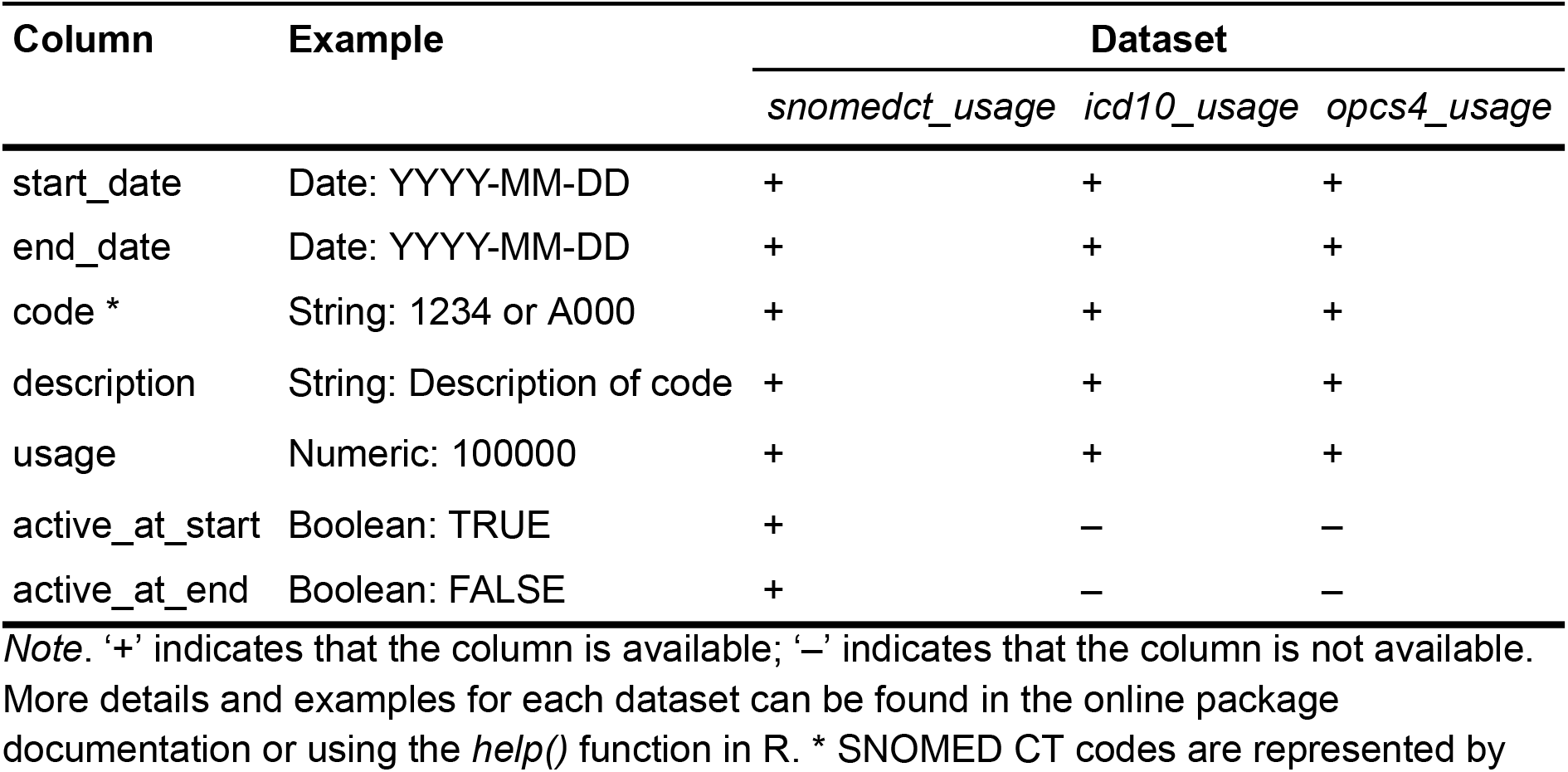
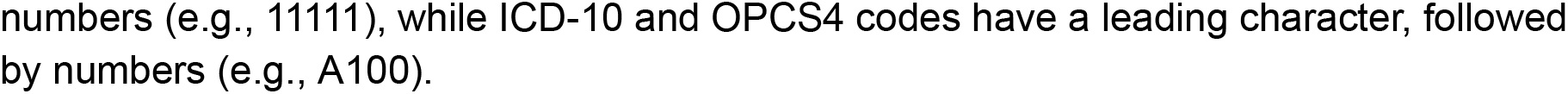
Available column names for each clinical coding system dataset in *opencodecounts* R package.

| Column | Example | Dataset |  |  |
| --- | --- | --- | --- | --- |
|  |  | *snomedct_usage* | *icd10_usage* | *opcs4_usage* |
| start_date | Date: YYYY-MM-DD | + | + | + |
| end_date | Date: YYYY-MM-DD | + | + | + |
| code * | String: 1234 or A000 | + | + | + |
| description | String: Description of code | + | + | + |
| usage | Numeric: 100000 | + | + | + |
| active_at_start | Boolean: TRUE | + | – | – |
| active_at_end | Boolean: FALSE | + | – | – |

*Note.* ‘+’ indicates that the column is available; ‘–’ indicates that the column is not available. More details and examples for each dataset can be found in the online package documentation or using the *help()* function in R. \* SNOMED CT codes are represented by
numbers (e.g., 11111), while ICD-10 and OPCS4 codes have a leading character, followed by numbers (e.g., A100).

#### Interactive online tool

We developed an interactive online tool - a Shiny app supported by the *opencodecounts* R package described below. The interactive [Shiny app](https://shiny.posit.co/r/reference/shiny/) visualises the data in the *opencodecounts* package and lets users explore trends over time for a selected set of codes. It can be launched locally with the *run_app()* function or accessed online using the *Launch Shiny App* tab at the top of the page on https://bennettoxford.github.io/opencodecounts/. The tool has a sidebar to select one of the available datasets (e.g., SNOMED CT, ICD-10, or OPCS-4) and search for a specific code or a collection of codes, including codelists from OpenCodelists.org.

Users can explore visualisations summarising overall trends and inspect individual code usage over time. A table provides key statistics on the frequency and proportional contribution of selected codes across all years. Additionally, the tool presents a structured list of all selected codes along with their descriptions, which can be copied or downloaded as a CSV file for use in EHR research or for upload to OpenCodelists.org.

#### R package: *opencodecounts*

Table 2 provides a summary of datasets and functions available to the users. One of these is a function *run_app()* which requires no arguments and launches the interactive tool described above. The current version of the package consists of two broad components: (1) three standardised datasets containing code usage counts for different clinical coding systems, and (2) a function to import specific codelists from OpenCodelists.org into R and the accompanying interactive tool (*Shiny* app).

**Table 2.**
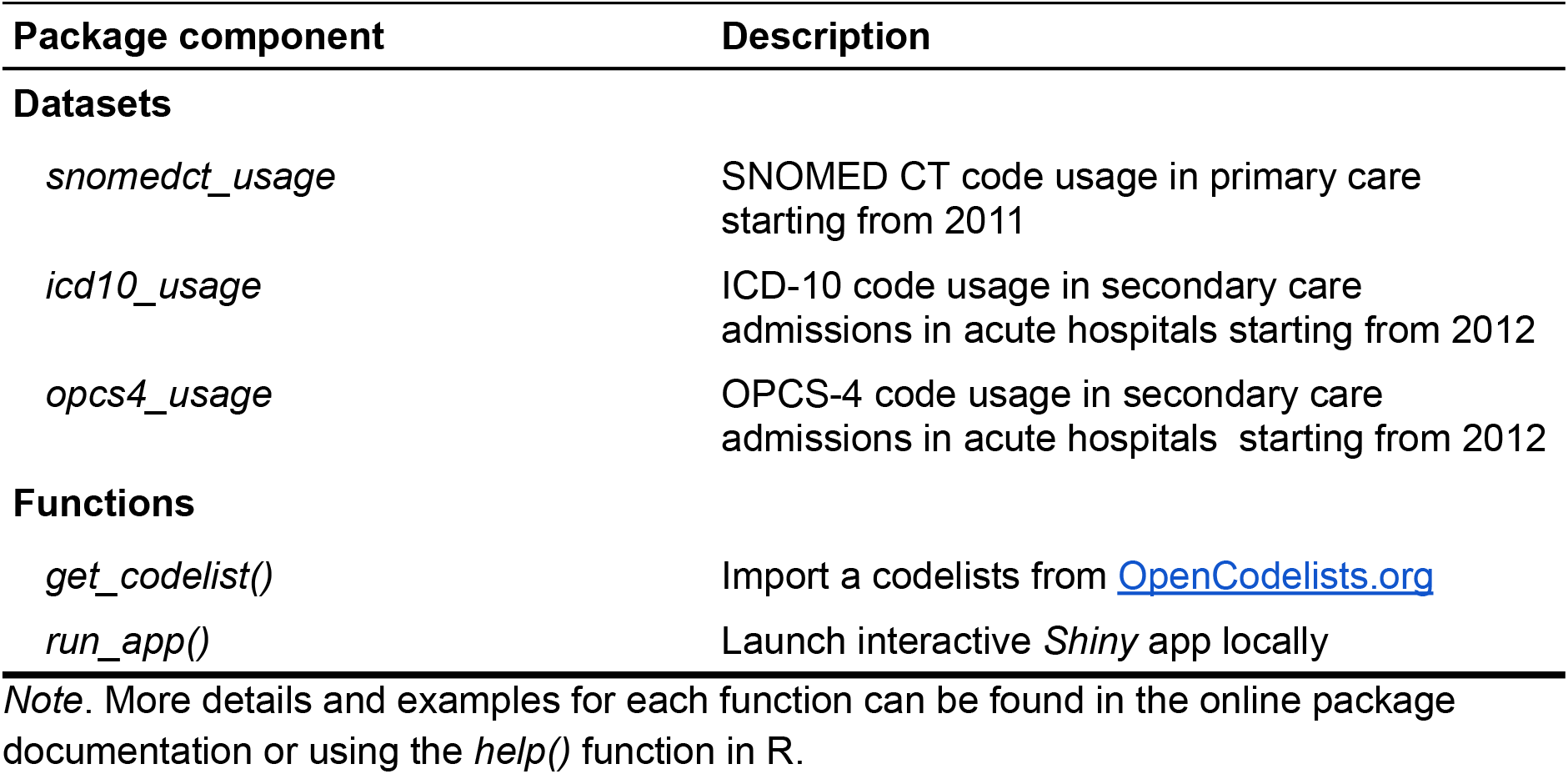
Main functions of the *opencodecounts* R package.

### Operation

The *opencodecounts* R package is freely available from Github and can be installed in R using *remotes::install_github(“bennettoxford/opencodecounts”)* and loaded using *library(opencodecounts)*. It requires an installation of R Version 4.1.0 or higher on Linux, Mac and Windows. The online tool is available using the *Launch Shiny App* tab at the top of the page on https://bennettoxford.github.io/opencodecounts/.

### Data and software availability and licensing statement

The package contains public sector information licensed under [the Open Government Licence v3.0](https://www.nationalarchives.gov.uk/doc/open-government-licence/version/3/). These data are publicly available from the NHS websites: SNOMED CT and ICD-10 & OPCS-4. The code for the software package and the *Shiny* app are available in the Github repository, under MIT licence.

### Use cases

We present several use cases that illustrate the application of *OpenCodeCounts*: (1) new codelist curation, (2) examining the feasibility of a research question, and (3) bespoke analyses in R. These examples are illustrative rather than realistic research examples. For further use cases and more detailed guidance, please refer to the R package documentation at https://bennettoxford.github.io/opencodecounts/.

Overall, the R package and the online tool summarise 41.7 billion instances of SNOMED CT code recording in primary care, and 1.3 billion and 400 million instances of ICD-10 and OPCS-4 code recording, respectively, in hospital admissions. The information on the frequency of usage is provided for 215,267 SNOMED CT, 12,380 ICD-10 and 9,857 OPCS-4 codes.

### Use case 1: Codelist development

Identifying code usage for all codes with a specific term can be very useful in codelist development. It provides an overview of the available codes for a specific clinical area and how much they are used. In this example, we are interested in traumatic injuries and want to examine trends in code usage of all procedures involving a “fracture”. To do so, we select the OPCS-4 classification system and enter the term “fracture” in the description box and select the “Show individual codes” field, see Figure 1.

**Figure 1.**
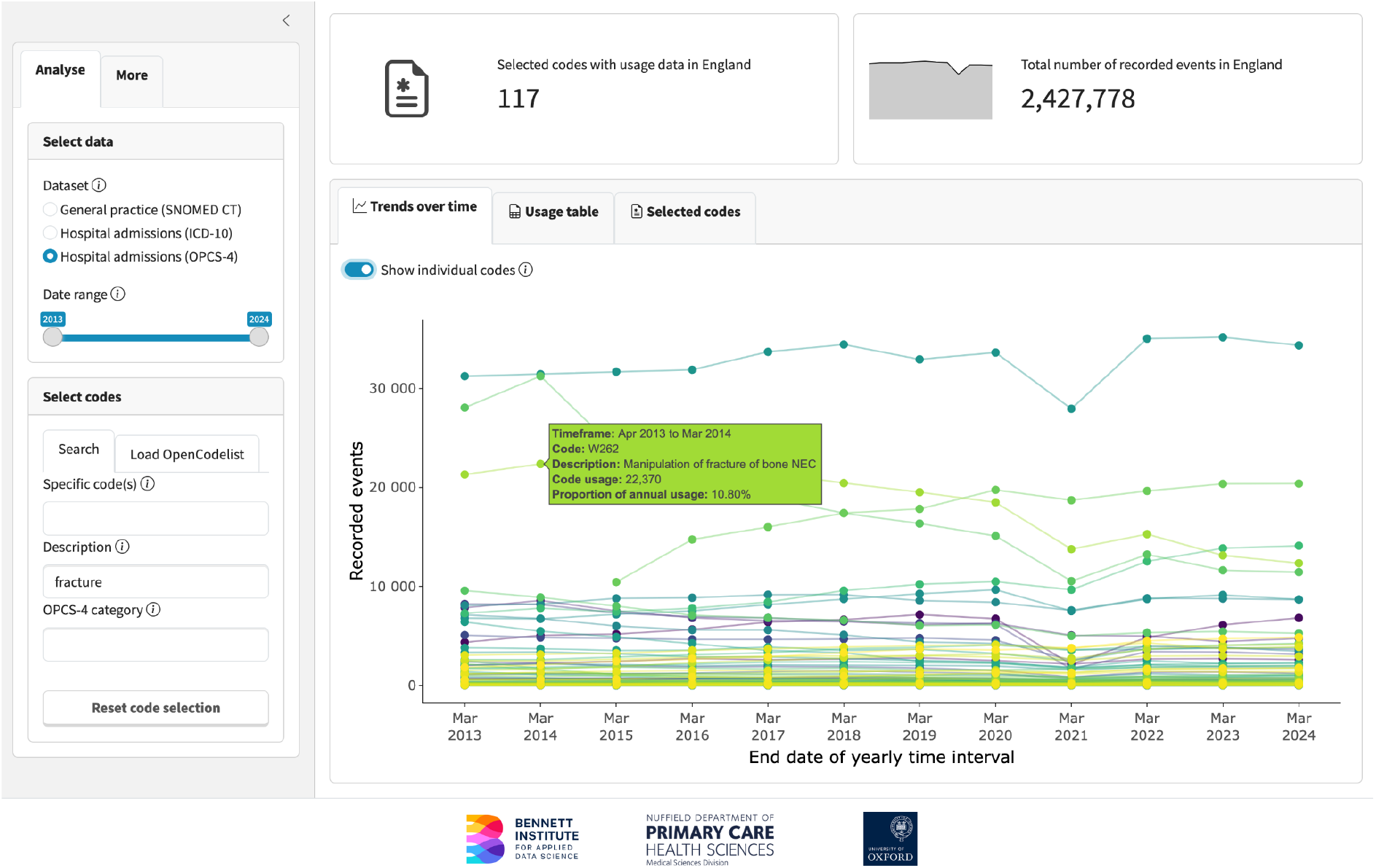
Screenshot from *OpenCodeCounts* online tool: exploration of individual code usage trends over time for all OPCS-4 codes with the term “fracture” in the description.

**Figure 2.**
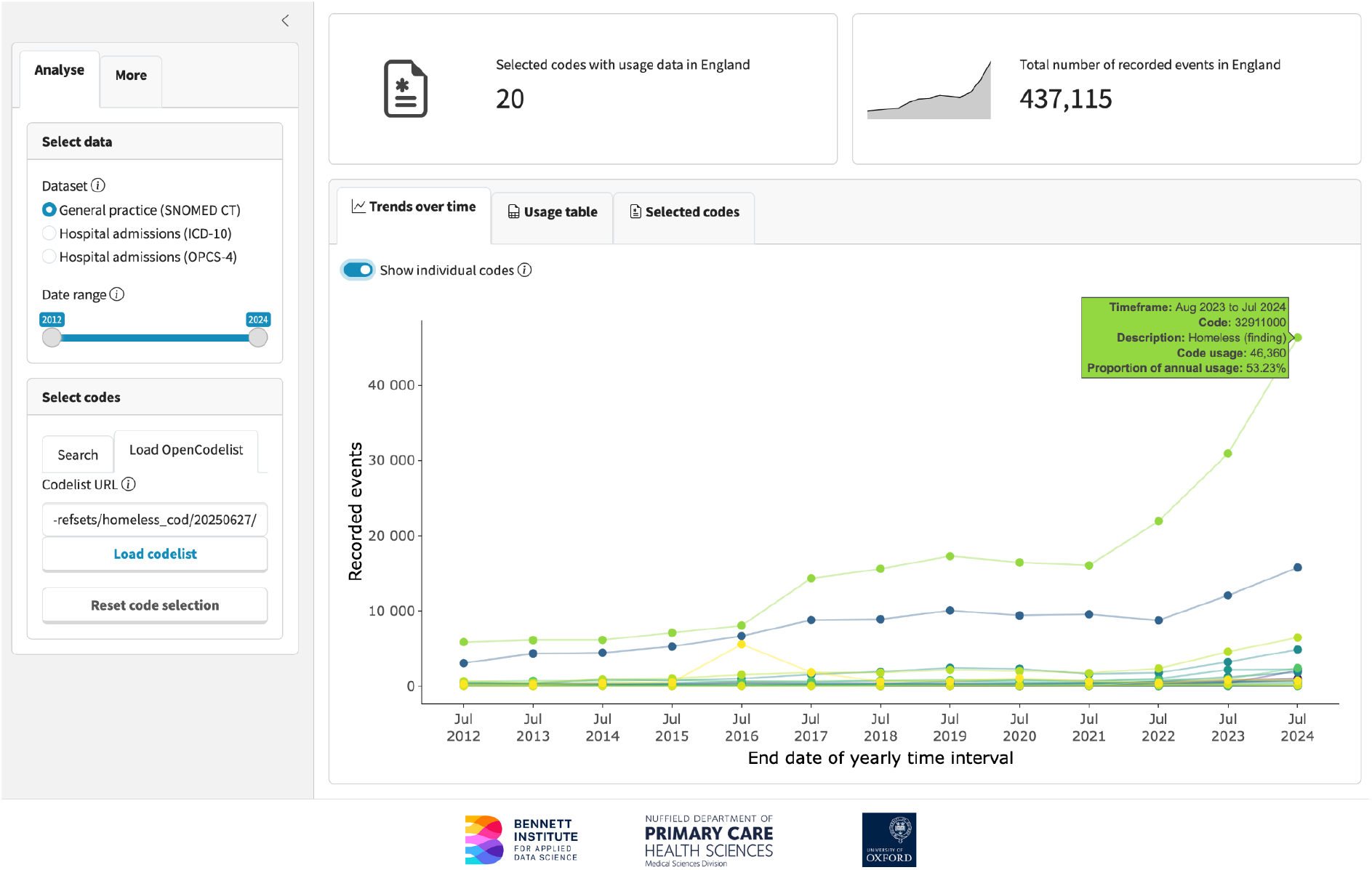
Exploration of code usage in the [“*Codes relating to homelessness*“](https://www.opencodelists.org/codelist/nhsd-primary-care-domain-refsets/homeless_cod/20250627/) codelist to estimate the feasibility of deriving a sufficiently sized cohort for EHR research.

This query returns 117 relevant codes with over 2.4 million recorded instances. We can display the exact code count, description, and annual proportion of code usage for each code by hovering over the corresponding data point. This can help to decide on the inclusion of edge cases (for example, fractures which may or may not be traumatic in origin) and identify the trends that may impact our study (for example, reduced coding of fractures during the COVID-19 pandemic). All codes containing the term “fracture” can be downloaded from the “Selected codes” tab as a.*csv* file and subsequently reviewed and used to create a codelist, for example via OpenCodelists.org.

### Use case 2: Assessment of study feasibility with England’s EHR

To assess study feasibility, researchers need to determine whether their variables of interest are sufficiently coded to facilitate their planned analysis. For example, EHR researchers interested in studying the health of people experiencing homelessness should check that SNOMED CT codes identifying homelessness are used frequently enough in primary care. For this example, we use an [existing NHS codelist](https://www.opencodelists.org/codelist/primis-covid19-vacc-uptake/homeless_cod/v2.5/), but we could also start by identifying codes using search terms, as described in the use case above. The analysis of the homelessness codelist shows around 80,000 recorded codes for homelessness in the financial year 2023/24. We compare these to the [official governmental statistics](https://www.gov.uk/government/collections/homelessness-statistics) (178,560 households assessed as homeless at application over four quarters of the year), and conclude that although EHRs may capture homelessness only in a subset of those who experience it, the coding is frequent enough to conduct a study using EHR data. The difference in numbers also highlights that individuals with homelessness recorded in primary care may differ from all those experiencing homelessness, which may affect our study interpretation.

### Use case 3: Bespoke analyses in R

The online tool allows users to select a custom group of codes, download the data and visualise trends. For bespoke analysis or visualisation, users can also work directly within R with access to the underlying data. This means users can produce visualisations and analysis for any of their chosen codes, across any available years and in a preferred graphical style. The *get_codelist()* function allows users to access codelists from OpenCodelists.org. The following examples demonstrate how to extract semantic tags from SNOMED CT codes (see Box 1) and how to find the three most used ICD-10 codes from a selected codelist and visualise their trends over time (see Box 2).

### Extracting semantic tags from SNOMED CT codes

Using the codelist for [Depression screening codes](https://www.opencodelists.org/codelist/nhsd-primary-care-domain-refsets/depscrn_cod/20250627/), we grouped code usage by semantic tag to explore how different tags were used over time, see Box 1. Figure 3 shows the yearly code usage across semantic tags such as “Procedure”, “Observable entity”, and “Finding”, highlighting an increased coding of observable entities (scores of depression screenings) in recent years.

**Figure 3.**
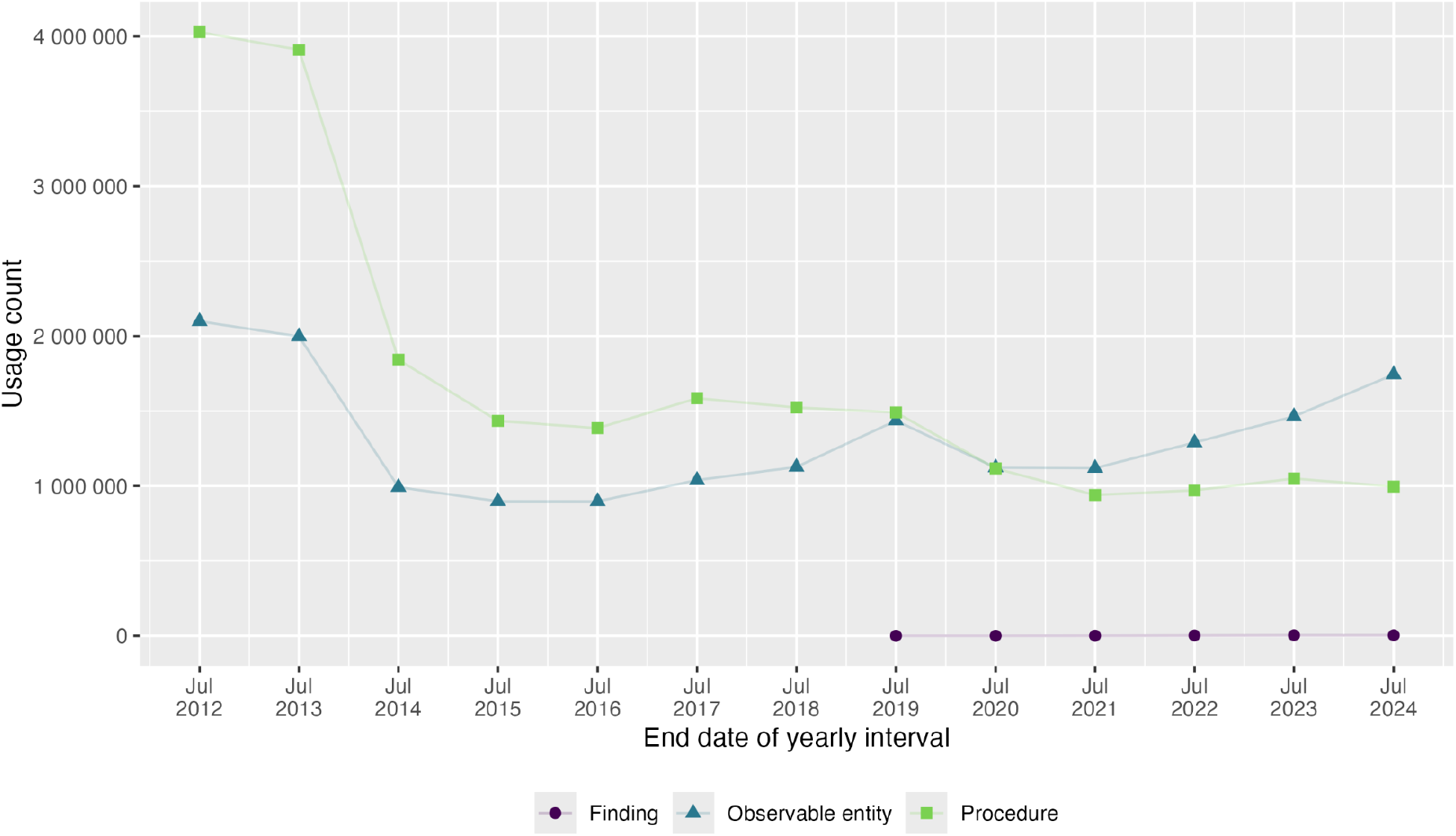
Trends over time in depression screening code usage grouped by semantic tag.

#### Box 1

**R code to extract and plot SNOMED CT semantic tags**

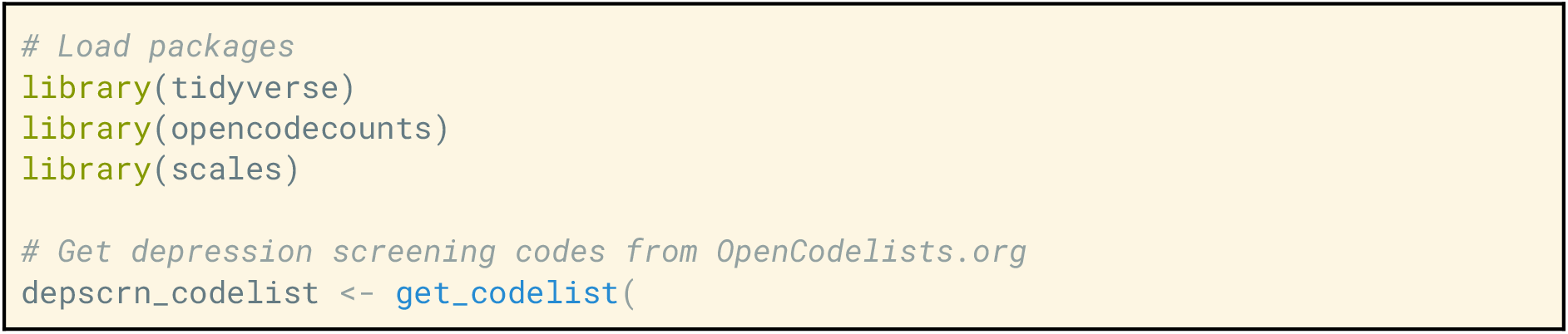
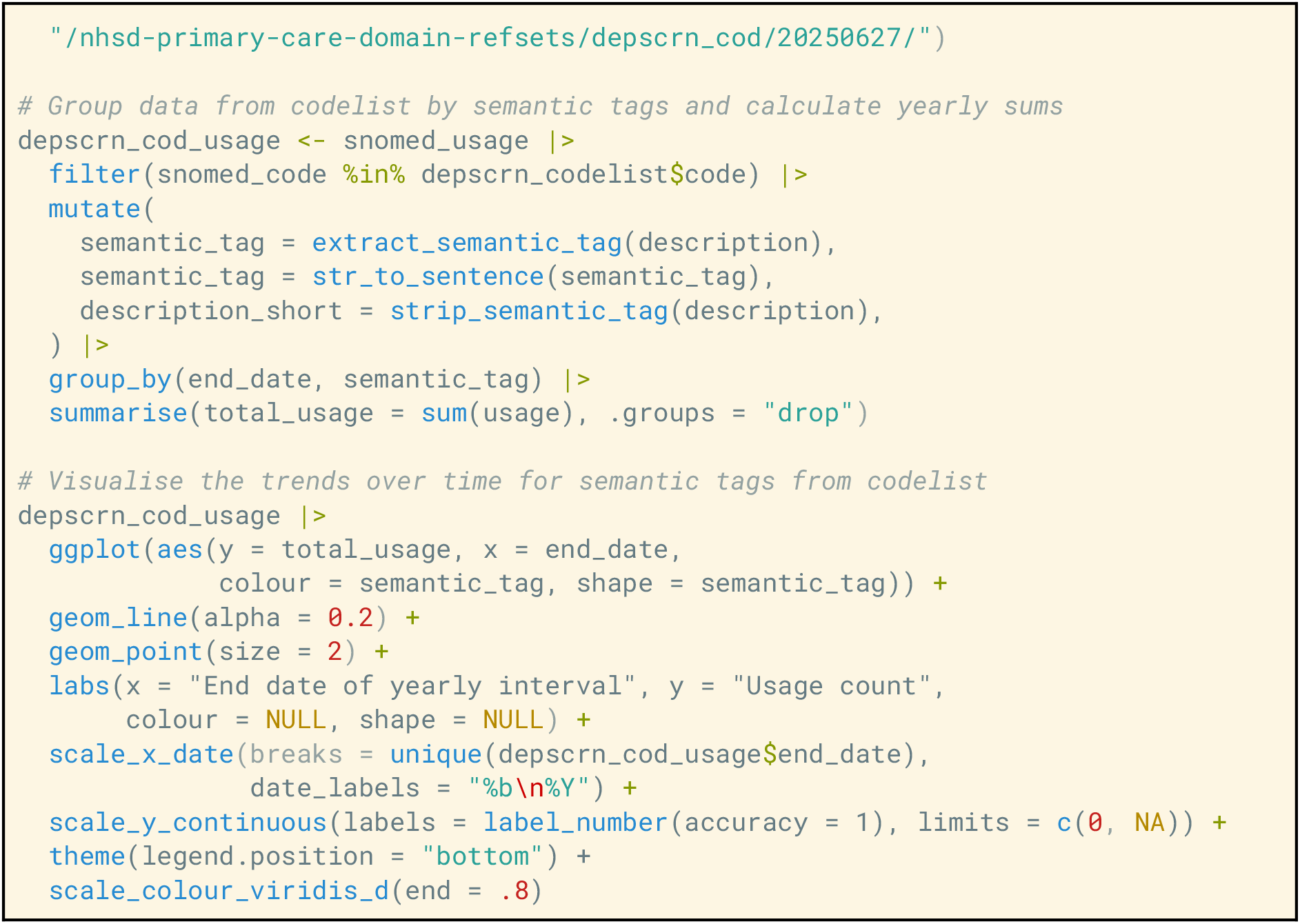

### Identifying the most frequently used ICD-10 codes

Using the ICD-10 Chapter XIX codelist (injury, poisoning, and certain other consequences of external causes), we identified the three most frequently used codes since 2014 and plotted their usage over time. The results highlight a relatively stable pattern for the most used code *Fracture of neck of femur* (S720), an increase in usage for *Poisoning: 4-Aminophenol derivatives* (T391) up until March 2020 and a decrease in code usage of *Superficial injury of other parts of head* (S008) from March 2022 onwards, see Figure 4.

**Figure 4.**
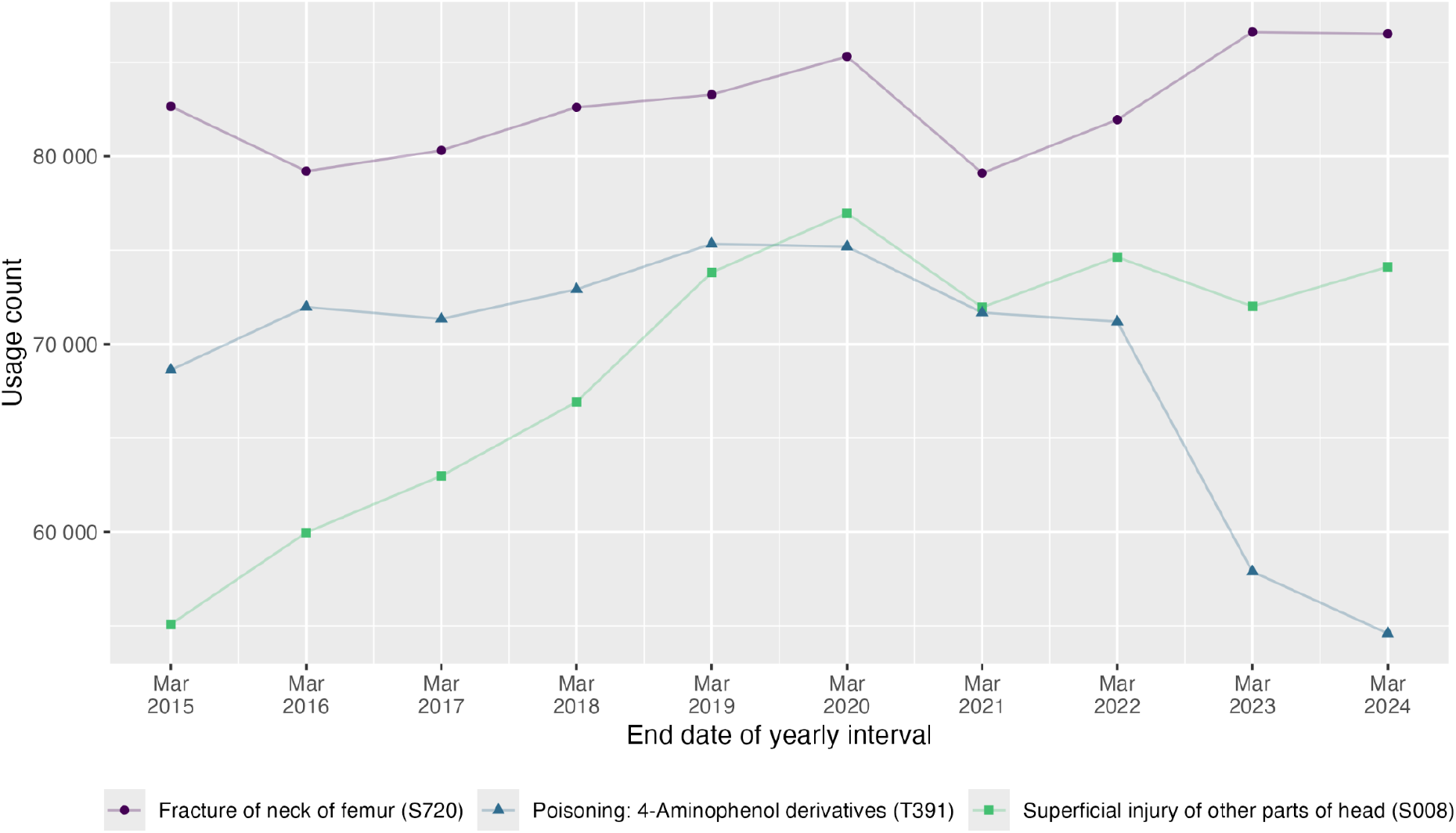
Trends over time in usage of the three most common ICD-10 Chapter XIX codes.

#### Box 2

**R code to identify and plot top three ICD-10 Chapter XIX codes**

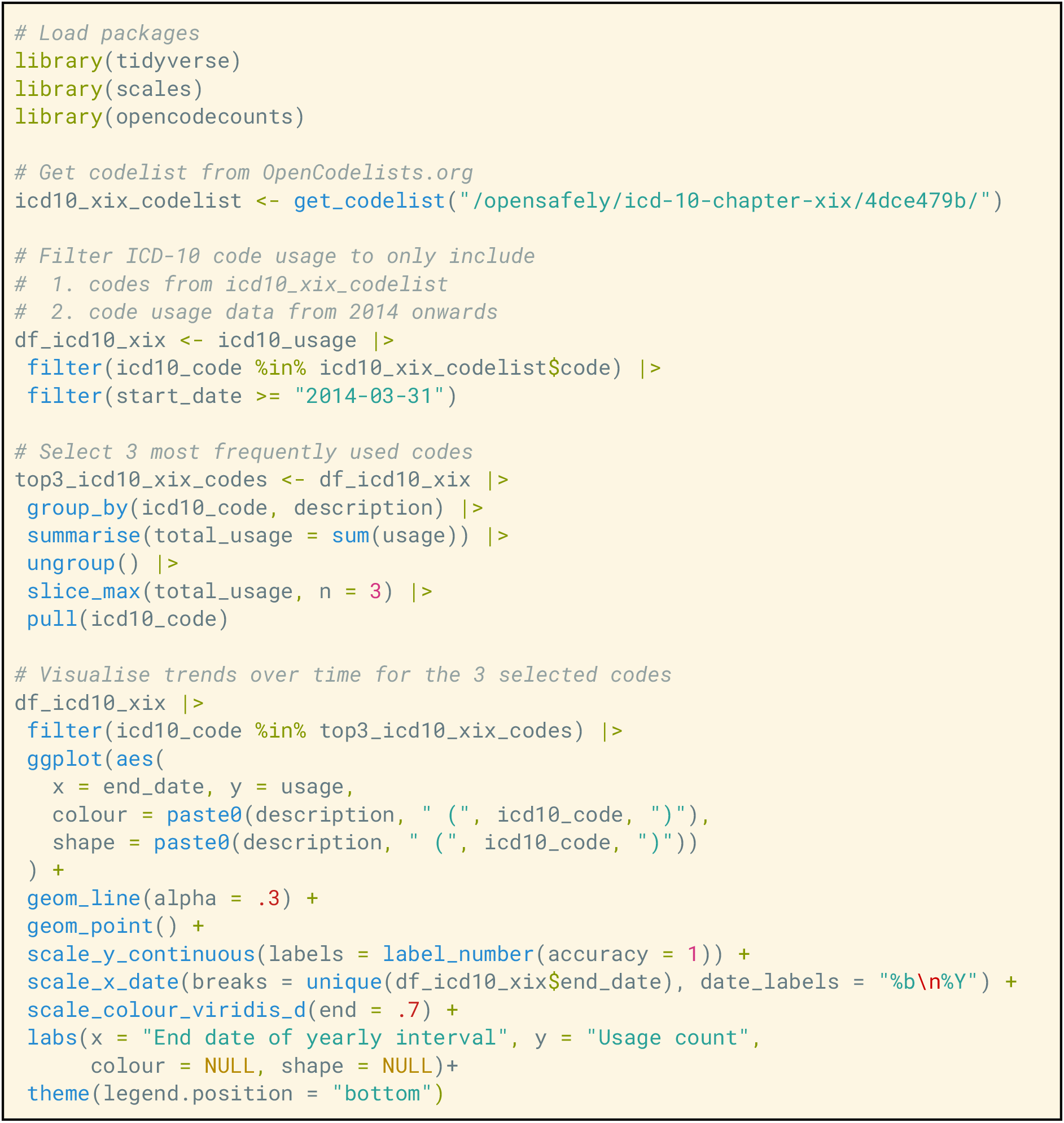

## Discussion

By making the national English clinical code usage data easily accessible, the *OpenCodeCounts* online tool and the accompanying *opencodecounts* R package are valuable tools for EHR researchers. They enable rapid exploration of trends in coding practices and the probability of deriving sufficiently sized cohorts and variables from routinely collected health records. They also aid codelist development, revision, and validation, and thanks to their compatibility with OpenCodelists.org, they promote codelist sharing and transparent curation. The combination of am online tool with an R package allows immediate and code-free data visualisation, without limiting the options for a bespoke, in-depth analysis in R.

A key consideration for using these tools is that the counts of clinical codes cannot be used to estimate disease incidence, service delivery statistics or the number of individuals with a certain characteristic. For example, OPCS-4 code usage counts should not be misinterpreted as the exact number of procedures conducted. This is because the surgical revisions may occasionally be recorded under the same code as the initial operation, and multiple codes can sometimes be recorded to make up one complete procedure. Similarly, the same diagnostic code resulting from one occurrence of an illness can be coded in several healthcare interactions, e.g. multiple GP appointments and/or hospital admissions, leading to multiple instances of code recording for a single disease case. For this reason, more complete electronic health data, with mapping between patient identifiers or service identifiers, is needed to accurately count service use or incidence. This is possible in trusted research environments such as [OpenSAFELY](https://www.opensafely.org/).

Another consideration for using these tools is that code usage data reflects both the underlying clinical activity and the external factors driving the recording practices. For example, the COVID-19 pandemic caused considerable disruption to the delivery of services across primary and secondary care, as well as to the clinical coding practices. Hospitals rely on good quality coding for reimbursement against the activity they undertake, in a system known as payment-by-results. This reimbursement scheme was temporarily suspended under the emergency conditions of the early pandemic and replaced by [block contracts](https://www.england.nhs.uk/coronavirus/documents/covid-19-block-contract-methodology/). This means that hospital reimbursement was uncoupled from coding for the financial year 2020, and thus the absolute coding of ICD-10 and OPCS-4 in the financial year 2020 does not reflect the volume of activity in hospitals. It is difficult to discern this from the genuine shift in hospital activity during the pandemic. Awareness of such factors is necessary to correctly interpret the data.

Similarly, financial reward programmes and mandatory reporting requirements can greatly impact the frequency of code recording. This means that any changes to these financial and regulatory incentives should be accounted for in the design of EHR studies. In primary care, a key driver of clinical coding is the Quality and Outcomes Framework (QOF). QOF consists of annually published business rules which define the clinical codes used for payments to practices. When an update to [QOF](https://digital.nhs.uk/data-and-information/data-tools-and-services/data-services/general-practice-data-hub/quality-outcomes-framework-qof) business rules changes which SNOMED CT codes qualify for payments, the uptake of the affected codes also changes radically. This means that codelists in clinical areas affected by QOF must be regularly reviewed to account for trends in code usage. Another example is the increasing use of SNOMED CT codes describing appointment characteristics since the 2019 introduction of mandatory reporting for the [NHS GP Appointments Data](https://digital.nhs.uk/data-and-information/publications/statistical/appointments-in-general-practice). Identifying when certain codes began to be used via the *OpenCodeCounts* can provide useful insights into study design.

Our tools only use a proportion of publicly available data. For example, OPCS-4 and ICD-10 code counts only use data from admitted patient care statistics, which do not capture hospital outpatient services. NHS publications also provide breakdowns by sex and age, which could be informative, but come at the cost of increasing the package size and reducing processing speeds. However, the constraints of including the data in an R package are balanced by making it readily available for analysis in an open-access environment and a language commonly used for EHR research.

Clinical coding has been increasing over the years, and the reasons for that are currently unclear. Patient-level analysis, in a secure data environment, would help examine whether this is driven by changes in healthcare utilisation and clinical activity, changes in coding practices or a mix of both. It may be that automated coding, a rise in online consultations, or the use of EHR templates increases the frequency of coding. A dedicated study in OpenSAFELY could examine the implications of these changes in coding trends for EHR research findings over time.

Future tool updates will integrate the new annual NHS data releases and consider the inclusion of additional datasets, such as outpatient and accident & emergency services, as well as demographic breakdowns. The tool will be maintained and suggestions for improvements and bug reports can be submitted by creating an issue in the [Github repository](https://github.com/bennettoxford/opencodecounts/issues/81).

In summary, *OpenCodeCounts* is an openly accessible online tool and R package that allows for a rapid and robust exploration of clinical coding trends. It captures over ten years’ worth of data across nearly all NHS-funded GP practices and hospital admissions in England. It provides an invaluable tool for EHR researchers, assisting in feasibility assessment and study design, codelist development and validation and results interpretation. The data presented should be interpreted with caution, as raw code counts do not equal the disease incidence or healthcare provision rates. For this, the analysis of individual patient data is required. *OpenCodeCounts* helps to prepare for such analysis and complements the reproducible EHR research ecosystem of OpenSAFELY and OpenCodelists.org.

## Data Availability

The data are publicly available from the NHS websites: SNOMED CT (https://digital.nhs.uk/data-and-information/publications/statistical/mi-snomed-code-usage-in-primary-care) and ICD-10 & OPCS-4 (https://digital.nhs.uk/data-and-information/publications/statistical/hospital-admitted-patient-care-activity) under the Open Government Licence v3.0. The code for the software package and the Shiny app are available in the Github repository (https://github.com/bennettoxford/opencodecounts), under MIT licence.

https://digital.nhs.uk/data-and-information/publications/statistical/hospital-admitted-patient-care-activit

https://github.com/bennettoxford/opencodecounts

## Authors contributions

Conceptualization: AAT, RH, BMK, HJC, LF, MW.

Data curation: AAT, VK, LO, KO, LF, MW.

Formal Analysis: AAT, YB, MW.

Funding acquisition: RH, AS, AG, WH, BMK, HJC.

Methodology: AAT, RH, BMK, HJC, LF, MW.

Codelists (subset of methodology): RH, YB.

Project administration: AAT, RH, MW.

Software: AAT, RH, VK, LO, KO, LF, MW.

Supervision: HJC, MW

Validation: AAT, RH, YB, VK, AS, WH, BMK, HJC, MW

Visualization: AAT, RH, MW

Writing – original draft: AAT, YB, MW

Writing – review & editing: AAT, RH, YB, VK, LO, KO, JM, AS, AG, WH, BMK, HJC, LF, MW

